# A Meta-Analysis of Influenza Vaccination Following Correspondence: Considerations for COVID-19

**DOI:** 10.1101/2021.06.10.21258685

**Authors:** Robert P. Murphy, Carol Taaffe, Elayne Ahern, Grace McMahon, Orla Muldoon

## Abstract

**Background:** High vaccination rates are needed to protect against influenza and to end the COVID-19 pandemic. Health authorities need to know if supplementing mass communications with direct correspondence to the community would increase uptake.

**Objectives:** The primary objective is to determine if sending a single written message directly to individuals increases influenza vaccine uptake, and a secondary objective is to identify any identified content shown to increase influenza vaccine uptake.

**Methods:** PubMed, PsycInfo and Web of Science were searched for English language RCTs testing a single correspondence for members of the community in OECD countries to obtain influenza vaccination. A meta-analysis with inverse-variance, random-effects modelling was used to estimate a mean, weighted risk ratio effect size measure of vaccine uptake. Studies were quality assessed and analysis was undertaken to account for potential publication bias.

**Results:** Twenty-two randomized controlled trials were included covering 37 interventions. Of the 37 interventions, 32 (86%) report an increase in influenza vaccination rates. A formal meta-analysis shows that sending a single written message increases influenza vaccine uptake by 18% (RR = 1.18, 95%CI [1.13-1.22], Z = 8.56, *p* < .001) relative to the no contact comparator group. Analysis shows that the intervention is effective across correspondence type, age group, time, and location, and after allowing for risk of publication bias.

**Limitations:** The review was restricted to English language publications, and the generalizability of results across the OECD may be questioned.

**Conclusions and implications:** The implication for public health authorities organizing vaccination programs for influenza, and arguably also for COVID-19, is that sending written vaccination correspondence to members of the community is likely to increase uptake.

The review was not registered nor was a protocol prepared due to time sensitivity.

---

Mass vaccination has a vital role to play in ending the COVID-19 pandemic. Given higher transmissibility of new variants, and an optimistic estimate of efficacy across the available vaccines of .80, achieving herd immunity requires a high rate of uptake of available vaccines.^1, 2^ When examining global trends in vaccination from 2015-2019, confidence in vaccination was identified as a key driver of improved vaccine uptake.^3^ As such, across many countries, public health authorities are using mass communications to address public confidence in the perceived safety, effectiveness, and importance of vaccination programs.^4-7^ Mass communications include public service announcements, media campaigns, notices to healthcare providers, and news coverage. A practical question for public health authorities, especially in OECD countries as they have well developed immunization programs and community wide access to social media, is whether supplementing mass communications with direct correspondence would increase vaccine uptake.

Previous experience in promoting adult influenza vaccination programs is relevant to answering this important question for COVID-19 vaccination programs. Crucially, both programs target the decision of adults on whether or not to vaccinate themselves. The transferability of learning about decisions from children’s vaccination programs to the uptake of COVID-19 vaccines by adults is limited, however, because concerns for dependent children can be very different to the concerns that parents and guardians have for themselves as vaccine recipients.^8^ Furthermore, it is plausible that perceived personal threat and risk with regard to age and health vulnerability to the adult influenza is patterned in the same way as COVID-19, thus highlighting the relevance of research examining influenza vaccination to the present pandemic.^9^ Examining the experience of influenza vaccination programs also has the advantage of providing results over four decades and covering a virus that was the cause of the most recent global pandemic prior to COVID-19: the 2009 pandemic hemagglutinin type 1 and neuraminidase type 1 (H1N1) influenza which had marked similarities to COVID-19 in terms of rapid spread.^10^

The aim of this systematic review and meta-analysis is to compile evidence from randomized control trials (RCTs) on the effectiveness of sending a single written message to an individual to encourage influenza vaccination. Our primary question is: “does sending a single written message directly to individuals increase influenza vaccine uptake?” Since the specific content and design elements of correspondence may play an important role in vaccine uptake, secondary questions are: “what content included in any correspondence is shown to increase influenza vaccine uptake?” and “is there any evidence of the comparative effectiveness of different content or design elements?” Several previous systematic reviews, listed in the methods section, examine methods for increasing influenza vaccine uptake but none of the previous reviews address our specific research questions.

## Methods

### Inclusion Criteria

Studies were included if they met the following criteria: compared influenza vaccination rates where a single correspondence was sent versus no correspondence; was a randomized controlled trial with an appropriate control group; was published in English in a peer-reviewed journal; was not specific to health care workers; and was conducted in an OECD country (given the particular relevance of such countries to the primary study question). The reference to “appropriate control group” refers to the fact that one study was excluded because it was reported as a randomized controlled trial with a control group but the control group was judged not appropriate because it comprised pregnant women who reported not participating in an ‘opt in’ SMS information service.^11^ Each record was screened by CT and the results independently reviewed by RM.

### Moderators

Information was recorded for each study on (a) the country in which it was conducted, (b) year of publication, (c) age group. We also classified interventions into (d) type of correspondence and (e) summary assessment of risk of bias. We attempted to identify if interventions were personalized or not, using the definition of a previous sytematic review^12^ of personalized communication as that “which aims to make a personally relevant appeal to individuals by, for example, using direct contact or individually addressed correspondence”, but insufficient detail was reported to classify all interventions (see Table A.1). Subgroup analysis was conducted based on the classification of (a) to (e).

### Search Strategies

A search was undertaken of Web of Science (all databases), PsycINFO (empirical studies) and PubMed in February 2021 using the search string below. CT and RM also hand-searched the references of eight systematic reviews found in the above search^12-19^, one meta-analysis^20^, and a rapid systematic review discovered through other means^21^.

The search string used was:

*((vaccine* OR *immunis*) AND (flu OR influenza) AND (letter* OR email* OR SMS OR text OR postcard* OR brochure* OR reminder* OR invitation* OR “portal message”) AND (vaccinated OR vaccination rate* OR uptake OR take-up OR effectiveness) AND (RCT OR trial OR quantitative OR experiment*))*

### Coding of Outcomes and Content

CT collected data on the outcome of interest: the effect of interventions on vaccine uptake. This data was reviewed by EA, with odds/ risk ratios calculated for the purposes of meta-analysis. Where data was missing, percentage vaccinations were collected. Study/ sample characteristics were also collected for use in subgroup analyses: year of study; country; patient type (e.g. chronic illness, elderly, healthcare insured); high risk group (Y/N); patient age group.

Content of messaging interventions was coded in part by RM and completed by CT, with each reviewing the other’s coding. Fourteen studies showing an effect included information on the content of correspondence; this content was coded into 18 elements, grouped as follows: (a) recommendation to get the vaccine; advice to get the vaccine soon; advice to get the vaccine every year; (b) information about the clinical manifestations of influenza; statement on the seriousness of influenza/ possible complications from influenza; statement that the vaccine helps avoid serious complications/ is effective; (c) statement that the vaccine is safe/ has minimal side effects; statement that the vaccine can cause minor side effects; addresses common concerns about the vaccine; (d) statement on the importance of the vaccine for high-risk people; statement of who is at high risk of complications from the flu; statement that the recipient is at high risk of complications/ a serious case of the flu; (e) information on how and where to get the vaccine/ scheduling information; access to online scheduling; clinic operating hours; clinic locations; information on the availability of the vaccine; statement that the vaccine is free. The template data collection forms and the data extracted from included studies is available upon request.

### Assessment of Risk of Bias

The Cochrane Risk of Bias tool was administered to assess the risk of bias across the studies included in the systematic review and meta-analysis.^22^ This tool consists of six bias domains assessed across seven items: selection bias (random sequence generation, allocation concealment), performance bias, detection bias, attrition bias, reporting bias, and other bias. A judgement of high, unclear, or low risk of bias was assigned based on the reported trial characteristics. Each study record was assessed by either EA or GMcM, with a sample (11/22; 50%) of records blindly and independently assessed by both EA and GMcM for reliability. Any conflicts between assessors were discussed in relation to the supporting information provided by the assessor for the bias judgement and a final consensus was agreed between EA and GMcM.

For the studies included in the meta-analysis (n = 21; k = 33) a summary assessment of risk of bias was computed using three of the domains from the risk assessment tool and studies were categorized into three groups based on the summary assessment using the framework as recommended by the Cochrane Risk of Bias tool: high, unclear, and low risk of bias.^22^ The three domains used were selection bias (concealment of allocation prior to randomization), performance bias (blinding of participants and study personnel), and detection bias (blinding of outcome assessors). This selection was informed by previous work that identified allocation concealment and blinding as the components of methodological quality most closely associated with the estimate of intervention effect.^23^ For example, inadequate concealment of allocation can introduce a bias if the investigator (and/or healthcare professional) has strong beliefs about the potential benefits of the intervention, which (in-)directly may confound the intervention process.

### Statistical Methods for Estimating Effect Size

The events of vaccination and total events (i.e., subsample size, inclusive of events and non-events) from the intervention and control groups were inputted into Review Manager v5.4 to generate risk ratio effect sizes. This was calculated as (SI / NI) / (SC / NC), where SI / NI = the number of ‘success’ events (vaccination) divided by the total events in the intervention group and SC / NC = the number of ‘success’ events (vaccination) divided by the total events in the control group. When only the percentage vaccination rate for both the intervention and control groups was reported, the absolute risk was derived from this percentage using the relevant denominator (i.e., subsample size of the intervention group or control group) reported in the respective study.^24^

Inverse-variance weighted, random-effects modelling was conducted to determine the mean risk ratio across the included studies. A random-effects model was selected to account for variability between studies which can likely be explained by factors other than sampling error^25^, for example, variance in the sample characteristics and the intervention components between studies. The risk ratio effect size contributed by each study was weighted by its inverse variance so that studies with a larger sample size were given more weight in the analyses to ensure precision in the mean, weighted effect size estimate.^25^ Each study contributed only one effect size to the meta-analysis per written correspondence intervention; this avoided weighting individual studies by the number of subsamples reported (e.g., if vaccination was reported by age group for the respective intervention) and also to ensure statistical independence of effect sizes.^26^

A mean, weighted effect size and 95% confidence intervals were generated for the meta-analysis and presented visually in a Forest plot along with the study-level effect sizes. The Z statistic was interpreted against a .05 alpha level to test the null hypothesis that the mean, weighted effect size was 0; a significant Z statistic indicated that the mean, weighted effect was significantly different from 0. Heterogeneity, resulting from differences between the study-level effect sizes that contributed to the mean, weighted estimate, was evaluated with the Q statistic Chi-square test. Due to low power in a meta-analysis with a small number of studies, the alpha level was set to .10, as recommended.^24^ The I2 index was applied to quantify the amount of heterogeneity between studies that could be explained by true heterogeneity rather than chance. This was interpreted in accordance with the recommended criteria: 25-49% = small, 50-74% = moderate, and 75%+ = large heterogeneity.^25^ [15]

Categorical variables such as the characteristics of the sample (age group), intervention (type of written correspondence) and study (location (continent), year of publication (decades), risk of bias assessment) were considered for subgroup analyses. A minimum of two studies were required per category in the subgroup analyses to ensure sufficient power to determine whether the categorical variable was a significant moderator of effect size.^25^

### Assessment of Risk of Publication Bias

A funnel plot (log risk ratio by standard error) was generated and visually inspected for asymmetry to determine the presence of publication bias. This is typically observed by missing studies towards the bottom of the graph on one side of the weighted, mean effect size line, indicating an absence of non-significant or unfavorable outcome studies with small sample sizes (publication bias). Egger’s test was conducted to quantify the funnel plot asymmetry and statistically determine the presence of publication bias. In the detection of publication bias, Duval and Tweedie’s trim and fill analyses were conducted to trim or remove extreme, positive, small studies and then impute the mirror of these studies to produce a symmetric plot and an unbiased, mean estimate of the intervention effect.^25^

## Results

### Sample of Studies

The full texts of 40 articles were screened for eligibility. A total of 22 randomized controlled trials were included in the review (see Figure 1). A description of the 37 interventions used in the studies is provided in the Appendix. One of the studies^27^ which accounted for 4 subsamples/intervention arms did not report the required statistics for inclusion in the meta-analysis so 21 studies (inclusive of 33 subsamples) were included in meta-analysis.

**Fig. 1.**
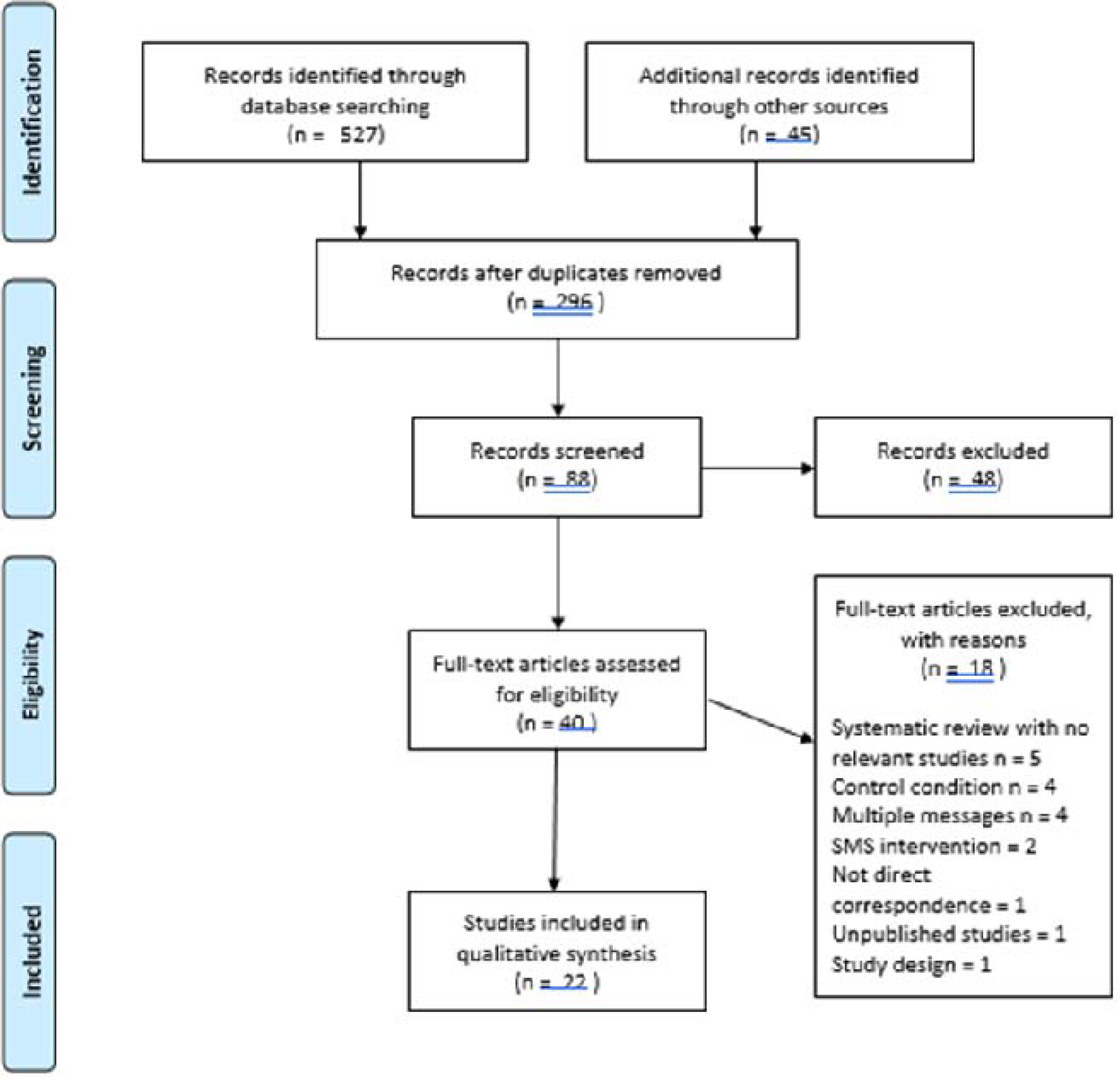
PRISMA Flow Diagram

### Study Characteristics

The studies were conducted in the USA, Canada, Spain, Denmark, New Zealand and Australia. Study populations were specified as at-risk or medical condition groups (n=9); older people (≥65 years) (n=7); Medicare beneficiaries (n=4); adults in the general population (n=1); and adults and children (>6 months) (n=1). Table A.1 provides a brief description of each intervention. A total of 37 types of broad intervention were used: letter (n = 23); postcard (n = 9); patient portal message (n = 2), educational brochure (n = 1), lottery (n = 1), brochure + lottery (n = 1). 25 interventions were characterized as “personalized”, with the remainder considered to be generic letters, postcards or portal messages.

### Risk of Bias

The quality assessment for the 22 studies revealed the lowest risk of bias for the first four domains: selection (random sequence generation and allocation concealment), performance, and detection bias. For each of these four domains less than 20% of the studies were judged to be of high risk of bias (details are available in the online supplemental file). However, for the first two of these domains, more than half of the studies were judged to be unclear due to the lack of transparency; selection domains of random sequence generation (11/22; 50%) and allocation concealment (14/22; 64%). The domain judged to have the lowest risk was detection bias, (14/22; 63%), reflecting blinding of participants and researchers; most studies measured outcomes with objective health records^28^ or insurance claim records^29^. The risk was judged to be high in more than half of the studies for the domains of reporting bias (14/22; 64%) -often only the percentage vaccination rate was reported without the corresponding frequencies of events and non-events -and attrition bias (12/22; 55%), due to the lack of explanation for attrition within some studies. High risk of other biases was noted in 41% (9/22) of studies. This was most often related to the possibility of sampling/recruitment bias. For instance, non-random sampling methods were often reported (e.g., site selection^30^) or the criteria used for exclusions may have limited the generalizability of findings (e.g., participant exclusion if believed to object to vaccination^31^).

As shown in Figure 2, two studies were deemed low risk across all seven domains ^28,32^, with a further seven studies judged as low risk in at least 4/7 domains ^29, 31, 33-37^. Although no study was deemed high risk across all domains, four studies were deemed high risk across at least 5/7 domains^6, 38-40^.

**Fig. 2.**
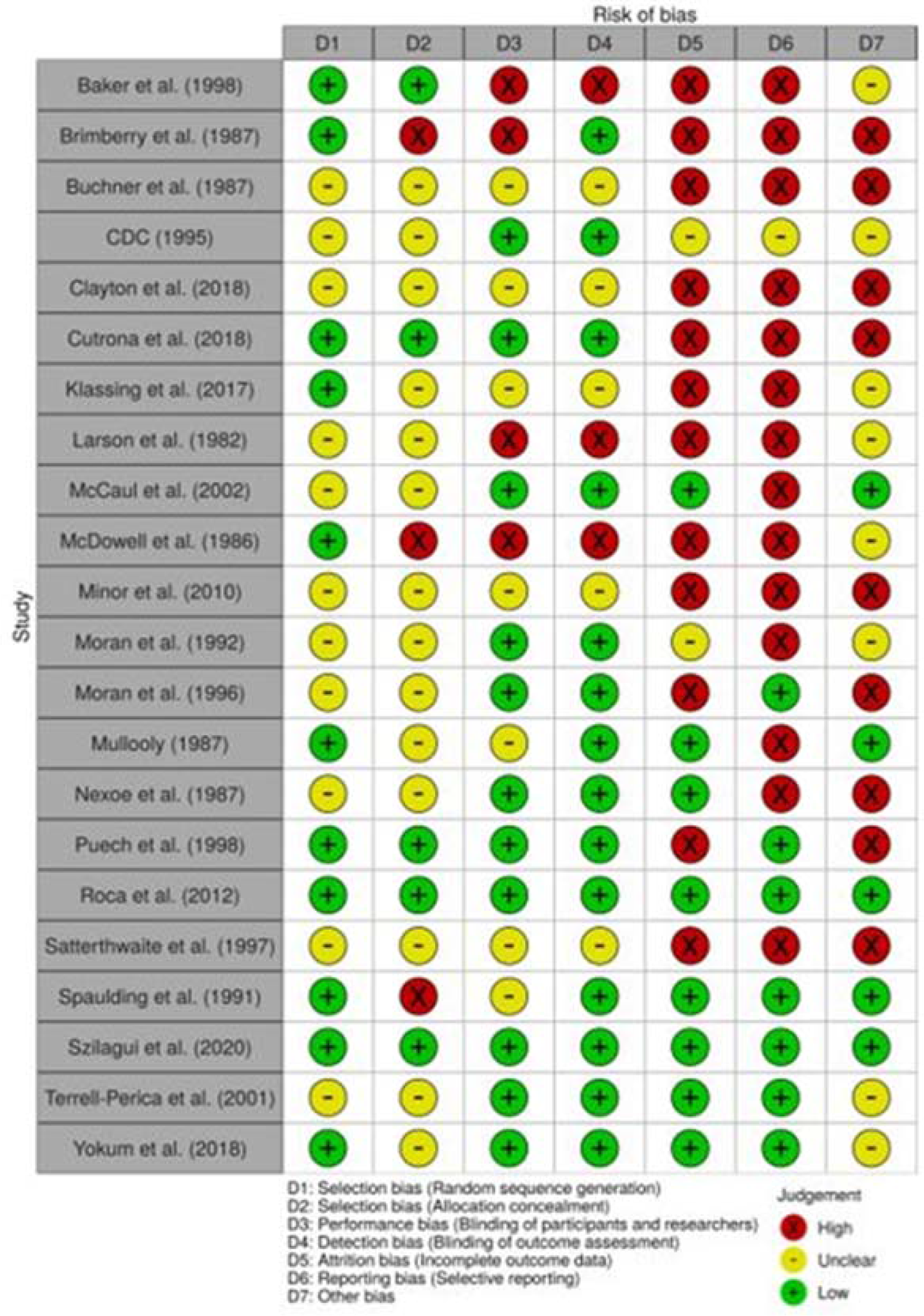
Traffic Light Plot: Risk of Bias Assessment

### Overall Effect of Correspondence in Each Study

Information on the reported effective size for each of the 22 studies is provided in Table 1. Of the 37 interventions, 32 (86%) are reported to have significantly increased influenza vaccination rates (i.e., where the odds ratio exceeds one or the p value is below 0.05). Two interventions showed no effect, and one showed an effect for men only. Sending a postcard to older people who had previously received a vaccine was not effective^41^. Combining an educational brochure with a financial incentive (a lottery to receive a gift certificate) was also not effective compared to sending either a brochure or the incentive alone^42^. A personalized postcard raised vaccination rates in men but not women and did not raise rates overall^31^. In two studies the intervention showed a negative effect on vaccination rates: in the first, pharmacists sent a personalized letter to asthma and COPD patients ^43^; in the second, a generic reminder letter slightly decreased vaccination rates compared to the control^39^.

**Table 1.**
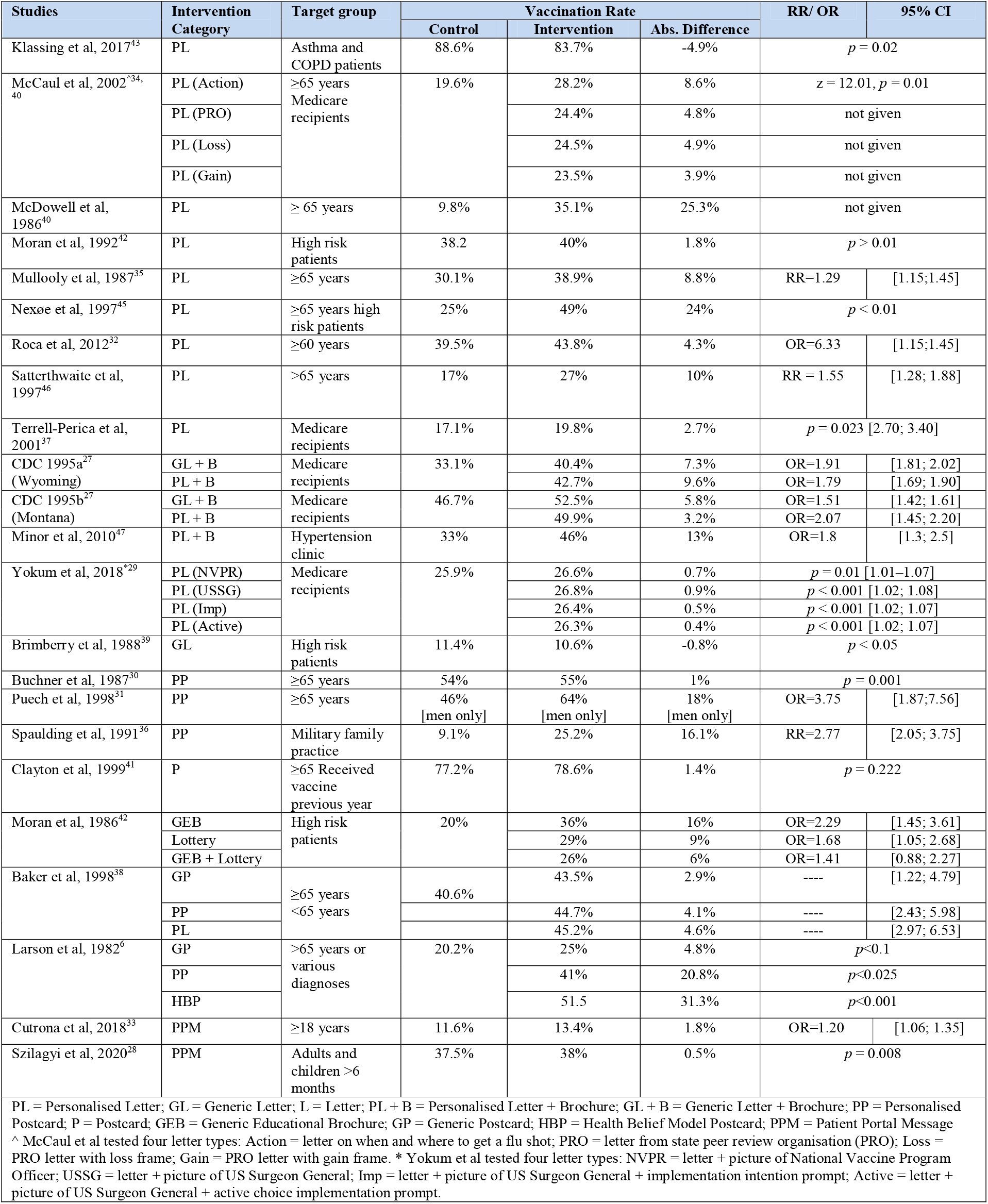
Intervention Type, Target Group and Effectiveness.

### Overall Estimates of Effect Size for Correspondence

The main analysis included 33 subsamples (intervention arms) across 21 studies (see Figure 3); one study did not report sufficient statistical information for inclusion in the meta-analysis^27^. Sending a single written message increased influenza vaccine uptake by 18%, relative to the no contact comparator group (RR = 1.18, 95%CI [1.13-1.22], Z = 8.56, *p* < .001). There was substantial heterogeneity among the included 33 samples (n = 21 studies), χ2 (32) = 390.95, *p* < .001, I2 = 92%) which warranted further subgroup analyses to determine the influence of patient and/or intervention characteristics on the effect size measure.

**Fig. 3.**
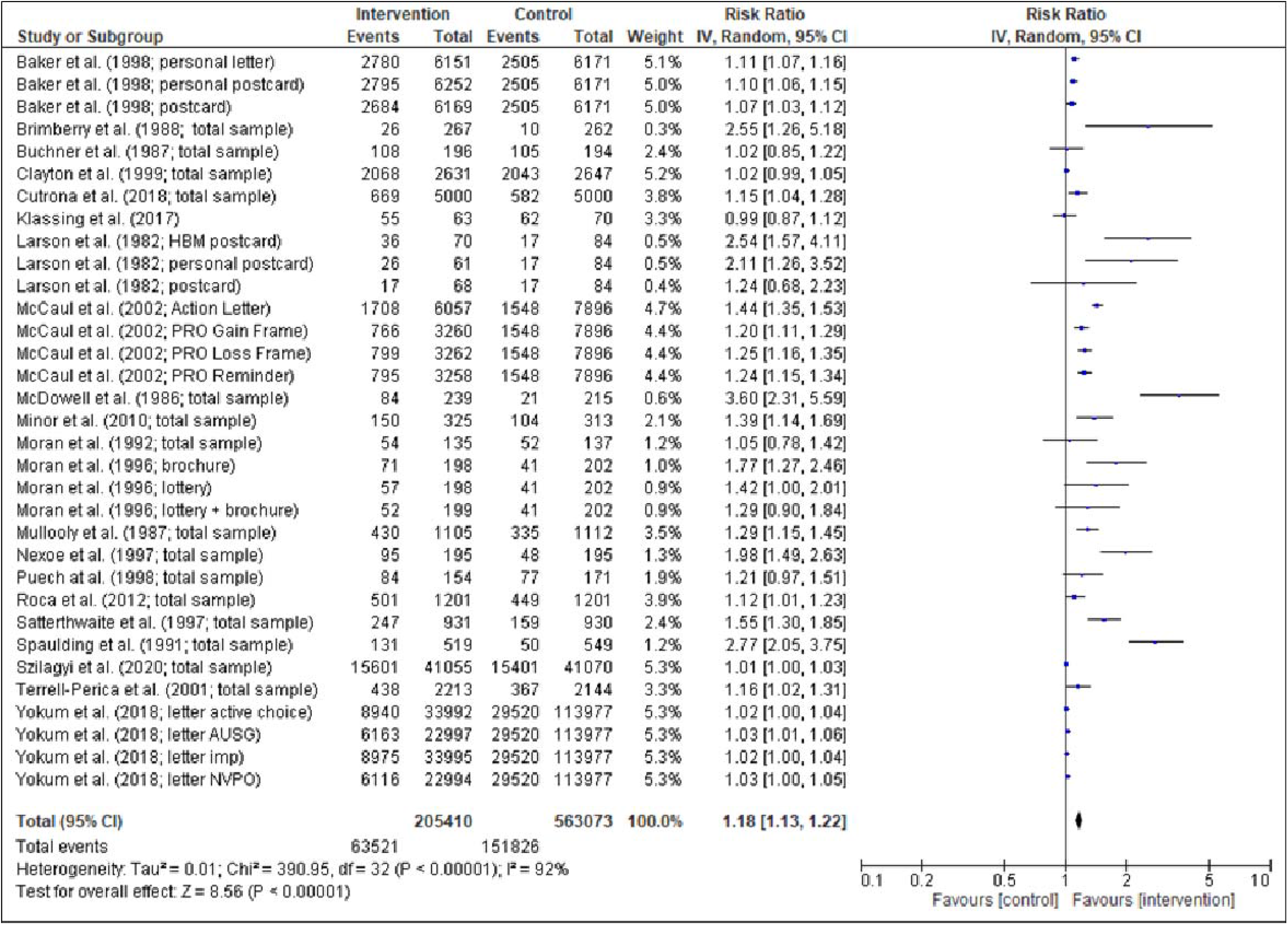
Overall Effect Size Estimate

### Subgroup Analyses

#### Type of correspondence

No significant differences were observed in the effectiveness of messaging based on the type of correspondence (letter, postcard, letter/postcard + brochure, portal message), χ2 (3) = 5.30, *p* = .15 (see Figure 4).

**Fig. 4.**
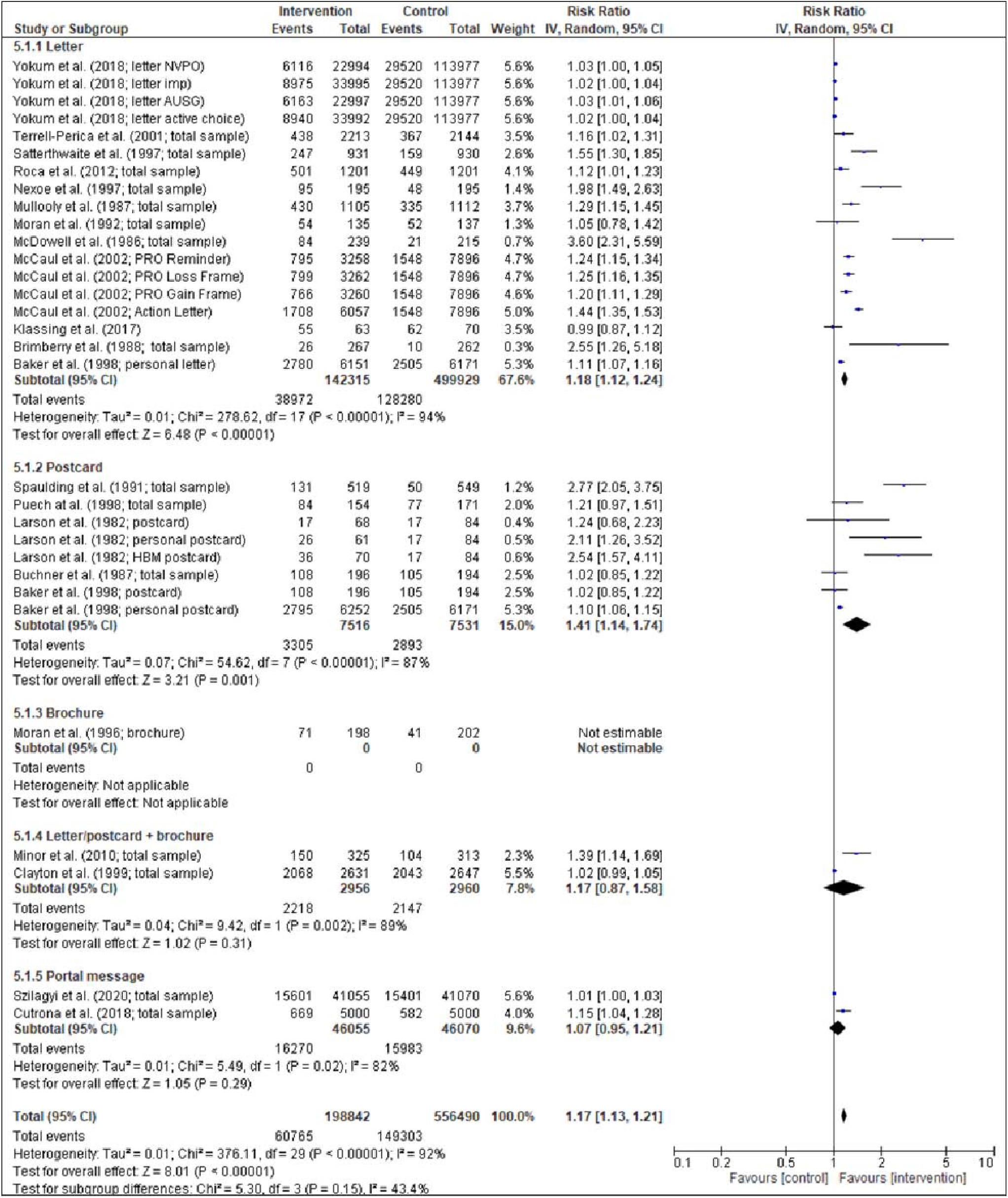
Effect Size Estimates by Correspondence Type

#### Continent

No significant differences were observed in the effectiveness of messaging based on study location (continent: North America, Europe, Australia), χ2 (2) = 2.59, *p* = .27 (see Figure 5).

**Fig. 5.**
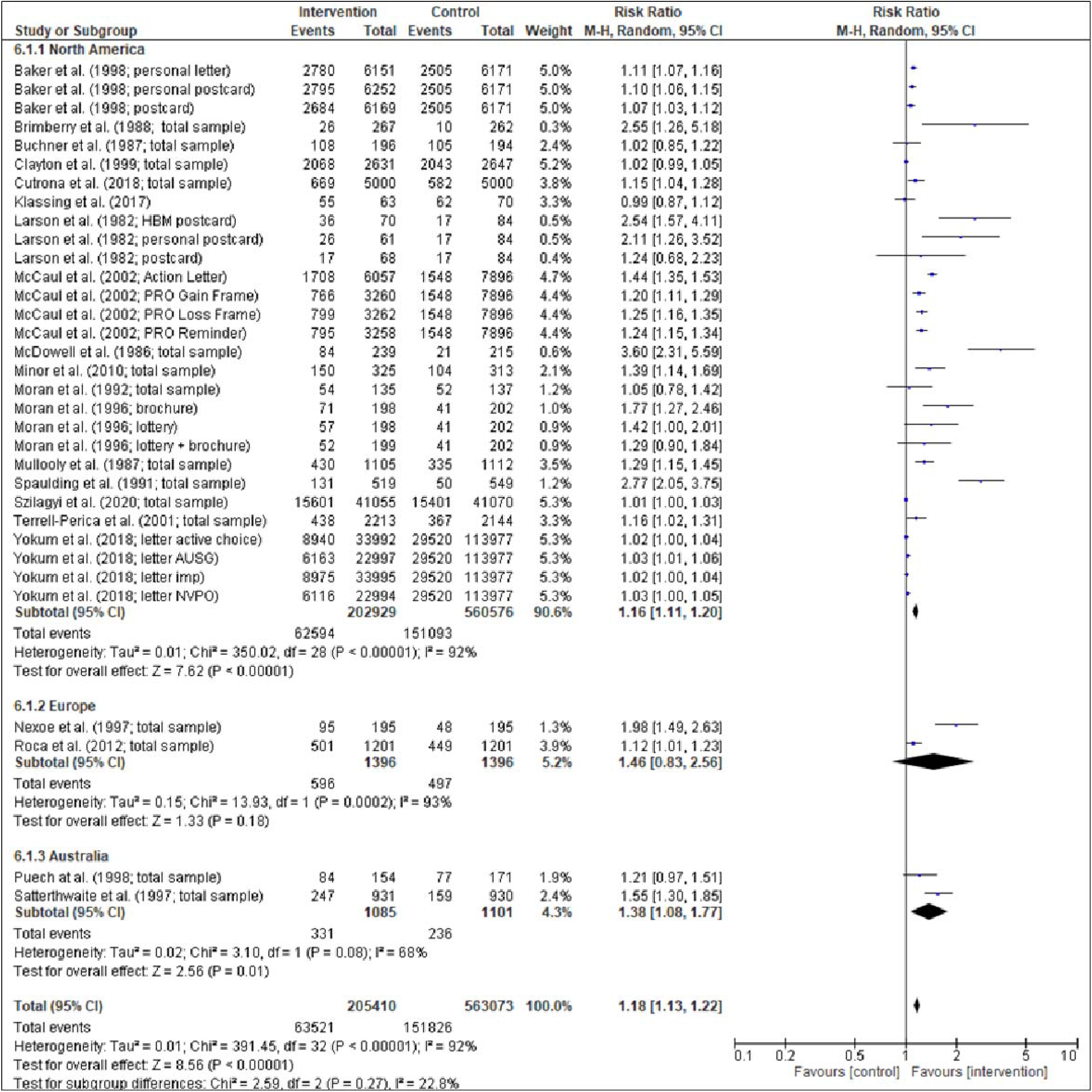
Effect Size Estimates by Location

#### Year of publication

Subgroup analysis by year of publication was carried out in two-decade intervals, i.e., 1980-1999 and 2000-2020. The effect of sending correspondence holds over both periods but was higher in the earlier period. Studies published in 1980-1999 saw a 33% increase on control (RR = 1.33, 95% CI [1.23, 1.44]) while the increase was 12% in those published from 2000-2020 (RR = 1.12, 95% CI [1.08, 1.17]), χ2 (1) = 14.40, *p* < .001 (see Figure 6).

**Fig. 6.**
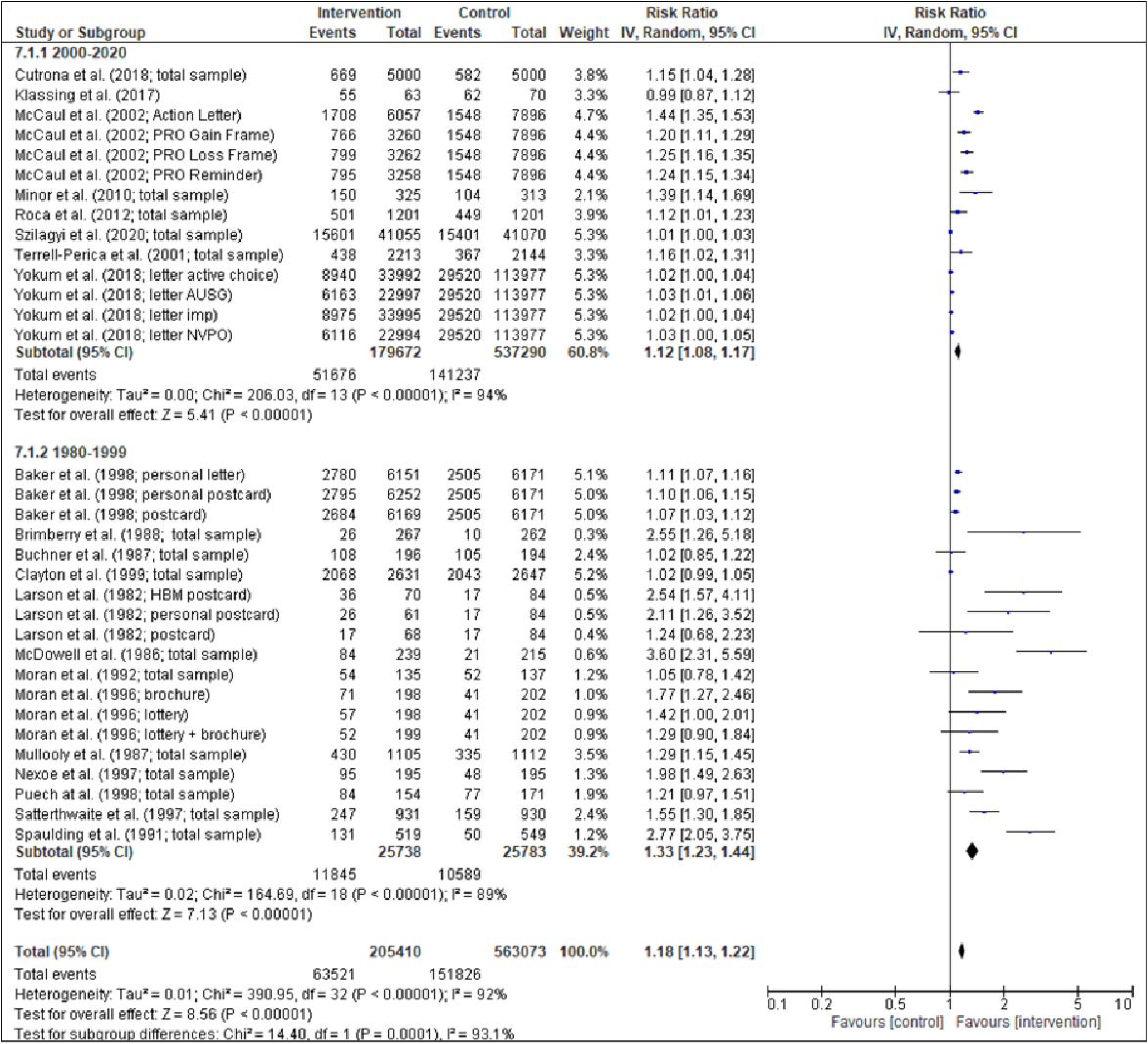
Effect Size Estimates by Year of Publication

#### Age group

A single direct message is shown to be effective across all age groups, but the size of the effect significantly differs, χ2 (1) = 3.21, *p* = .07. Following a message, the increase in uptake is greater (see Figure 7) for young and middle-aged adults (typically 18-64 years; 54% increase in vaccine uptake, relative to control) compared to older adults (typically ≥65 years; 16% increase in vaccine uptake, relative to control).

**Fig. 7.**
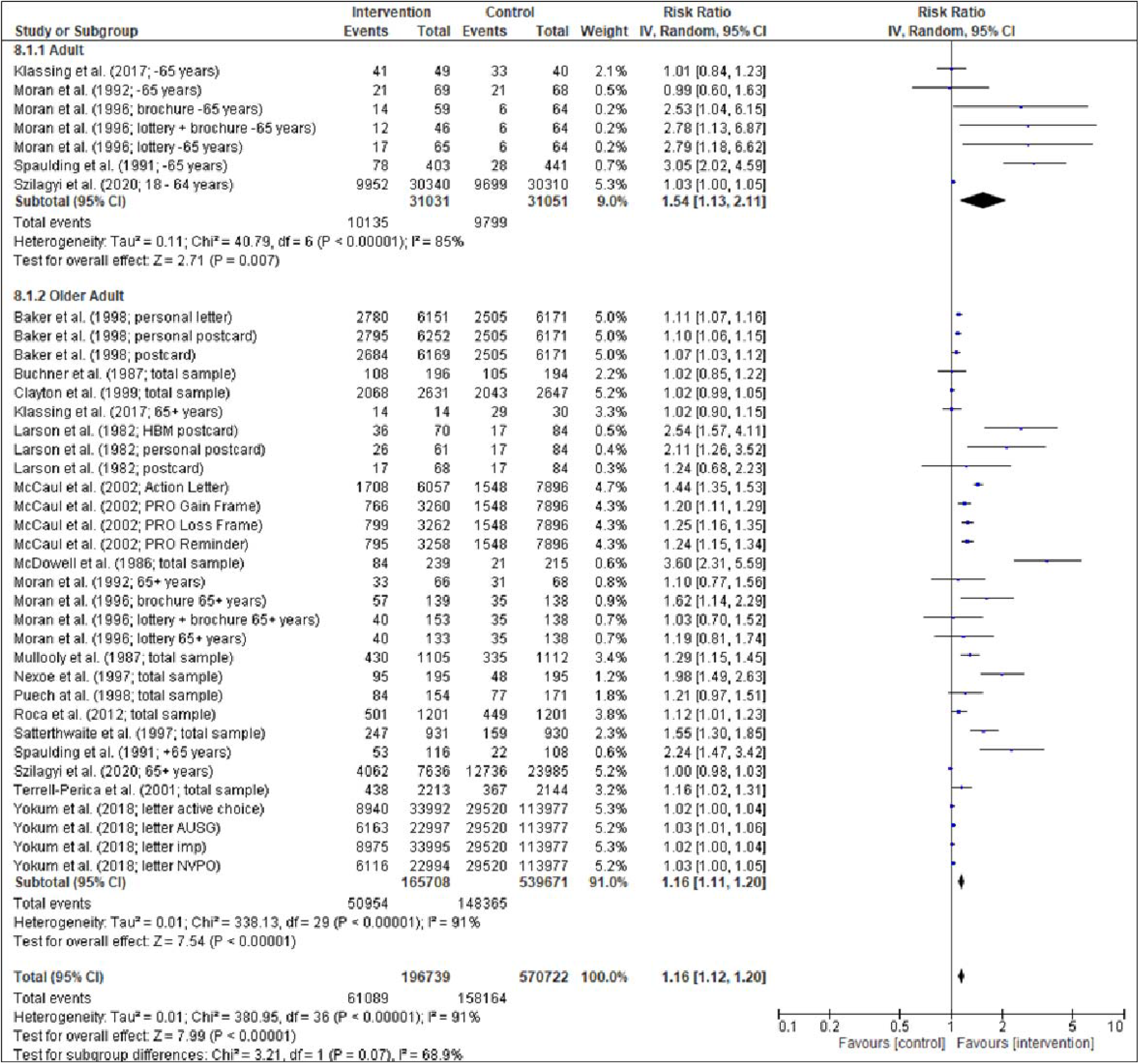
Effect Size Estimates by Age Group

#### Risk of bias assessment

Significant differences were observed in the overall effectiveness of messaging on vaccine uptake when risk of bias assessment was considered, χ2 (2) = 11.77, *p* = .003 (see Figure 8). In particular, the effectiveness of messaging differed significantly between low and high risk populations, χ2 (1) = 11.52, *p* = .001, unclear and high risk populations, χ2 (1) = 8.52, *p* = .004, but not between low and unclear risk populations, χ2 (1) = 1.69, *p* = .19. On average, messaging contributed to a 9% increase in vaccination (RR = 1.09, 95% CI[1.00, 1.19]) for low risk studies, a 17% increase in vaccination for unclear risk studies (RR = 1.17, 95% CI[1.11, 1.22]) and a 42% increase in vaccination (RR = 1.42, 95% CI[1.26, 1.61]) across high risk studies.

**Fig. 8.**
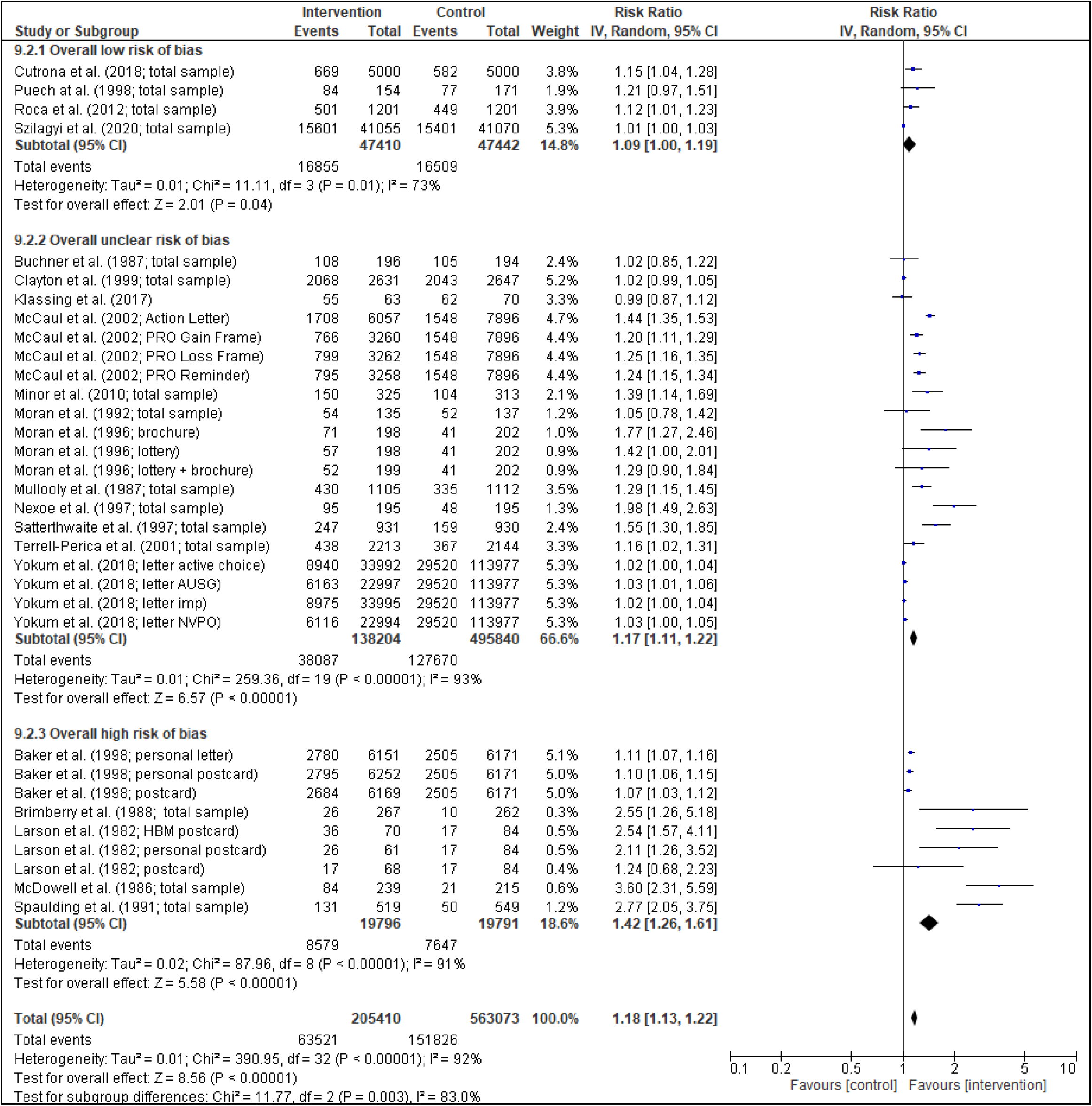
Effect Size Estimates by Summary Assessment of Risk of Bias

### Publication Bias

A visual representation of the publication bias via funnel plot (log risk ratio by standard error) was produced (available in the online supplemental file). Egger’s test was performed to quantity the funnel plot asymmetry and indicated that publication bias was present in the meta-analysis, with small sample studies with non-significant or smaller than average effect sizes likely to be missing (Egger’s intercept = 3.50, *p* < .001). Following trim and fill analyses to account for publication bias by imputing the effect sizes of 12, hypothetical, missing studies, the overall, mean, weighted effect size was adjusted to RR = 1.09 (95% CI [1.05, 1.13]). This corresponds to a 9% increase in vaccination following the messaging intervention, relative to control.

### Content Analysis of Correspondence

Of the studies with intervention arms showing an effect, 14 provided information on the content of correspondence: primarily a descriptive summary of the correspondence tested rather than the correspondence text in full. The most commonly reported content elements per study were: a recommendation to get the vaccine (n = 10); a statement that the vaccine helps avoid serious complications/ is effective (n = 7); a statement of the seriousness of the influenza/ possible complications from influenza (n = 7); information on how and where to get the vaccine/ scheduling information (n = 6); advice to get the vaccine every year (n = 5) and a statement that the vaccine is free (n = 5).

Other reported content elements were: a statement that the vaccine is safe/ has minimal side effects (n = 4); an address to common concerns about the vaccine (n = 4); a statement of who is at high risk of complications from the flu (n = 3); clinic operating hours (n = 2); clinic locations (n = 2); information on the availability of the vaccine (n = 2); a statement that the recipient is at high risk of complications/ a serious case of the flu (n = 2); statement on the importance of the vaccine for high-risk people (n = 2); statement that the vaccine can cause minor side effects (n = 1); advice to get the vaccine soon (n = 1); information about the clinical manifestations of the influenza (n = 1); access to online scheduling (n = 1).

### Difference in Effectiveness Across Intervention Arms

Four of six studies found a difference in results between intervention arms. The most effective interventions in these studies highlight design elements that might influence vaccine uptake. In one study the effectiveness of the intervention increased with more personal modes of contact: ‘the reminder postcard from the patient’s primary care physician was more effective than the generic postcard and the personalized tailored letter was more effective than either postcard intervention’^38^. Another study tested three postcard types and found that all were more effective than no reminder.^6^ A postcard designed according to the Health Belief Model was most effective (32.1% increase), followed by a personalized postcard (20.8% increase), while a ‘neutral’ reminder postcard showed a comparatively lower increase in vaccine uptake (4.8% increase). In testing four different letter designs, it was found that only the action letter (giving the exact time and places of vaccination clinics) was markedly more effective than the others: ‘First, differential framing was no more effective than providing a simple reminder. Second, providing action instructions had a powerful incremental effect on vaccination rates.^34^ An earlier study found that sending an educational brochure alone was more effective than either a financial incentive or sending both brochure and incentive: ‘the educational brochure more than doubled the likelihood of influenza immunization (odds ratio [OR] = 2.29, 95% confidence interval [CI] 1.45 to 3.61), whereas the incentive had less of an effect on immunization (OR = 1.68, 95% CI 1.05 to 2.68). Immunization for the group mailed both interventions was not significantly different from control.’^42^

Two of the six studies did not find a difference between intervention arms. One found no difference in sending a personalized versus a generic letter^27^: ‘The likelihood of vaccination was similar for persons who received a personal letter and for those who received a form letter.’ A study testing four letter types ‘found that a single mailed letter significantly increased influenza vaccination rates compared with no letter. However, there was no difference in vaccination rates across the four different letters tailored with behavioural science techniques.’^29^

## Discussion

The current review offers evidence from previous influenza vaccination programmes to inform future programmes targeted towards influenza and also the uptake of COVID-19 vaccination. The seasonal and consistent burden of influenza meant that infrastructure, prevention, and treatment strategies could be more promptly implemented in response to the 2009 pandemic, and this has not been the case in response to SARS-CoV-2.^10^ Nonetheless, we argue that there is important learning available.

First, our meta-analysis found that sending a single short correspondence to individuals increases the uptake of the influenza vaccine. The finding of an increase in uptake following direct correspondence holds across different types of correspondence (letters, postcards, letters/postcards + brochures, or portal messages), across different age groups (18-64 and 65+ years), across continents (North America, Europe, or Australia), and across time periods (1980-1999 and 2000-2020). The positive effect also holds after considering possible risk of study bias and potential publication bias.

While other reviews have examined methods to increase influenza vaccine uptake, this is the first study to examine and provide a meta-analysis of the effect of providing a single direct correspondence.^14^ Strengths of this review is that it exclusively included RCTs, followed the PRISMA statement, undertook quality assessment, and it accounted for the possibility of study and publication bias when estimating intervention effects. Weaknesses of this review include the restriction to English language publications. This review only included studies undertaken in OECD countries as these were felt to be most pertinent to the primary review question of whether supplementing mass communications with direct correspondence increases influenza vaccine uptake, as in these countries public health systems are well developed and most members of the community have access to mass media. The generalizability of results across the population of OECD countries may be questioned, as the studies were undertaken in six countries and 16 of the 21 studies in the meta-analysis related to older adults (≥60 years) or groups with specific medical conditions that might be considered at high risk from influenza.

These caveats aside, there is a second important implication for public health authorities organizing vaccination programs for influenza, and arguably also for COVID-19. Sending written vaccination correspondence directly to members of the community is likely to increase vaccine uptake more than using mass communications alone. When designing correspondence to support the uptake of the influenza vaccine, public health authorities should consider including the most reported content used in correspondence shown to increase influenza uptake. In particular, it is important to give a clear and strong recommendation to be vaccinated; provide information on vaccine effectiveness, the seriousness of influenza and how vaccination can avoid complications; state that the vaccine is safe; as well as providing information on cost and instructions on how and where to get vaccinated. These factors are also likely to be relevant for inclusion in correspondence to support the uptake of COVID-19 vaccines as they address many of the most frequently cited reasons by citizens in OECD countries for willingness and unwillingness to obtain COVID-19 vaccines, as identified in a recent review of peer reviewed papers.^44^ Based on the findings in the same review it would also be advisable for correspondence supporting the uptake of COVID-19 vaccines to briefly explain the speed at which COVID-19 vaccines were developed and tested, and for mass communications to support trust in health professionals, government agencies and in science.

Further research is needed on designing direct written communications to maximize vaccine uptake, whether in paper format or electronic media. In publishing results it is advised to quote the full text of tested correspondence to allow comparative analysis of effective design elements. To conclude, this meta-analysis provides evidence for single, direct messaging in increasing vaccination uptake for the influenza vaccine and can provide important insights for the rollout of vaccination programs for COVID-19.

## Data Availability

The template data collection forms and the data extracted from included studies is available upon request.

---

**Table A.1.** Summary of Interventions

| Studies | Interventions |
| --- | --- |
| Klassing et al, 2017<br>USA<br>n = 311 | Control: No contact<br>Intervention: (1) standardized letter, or (2) phone call. A phone call script was utilized for the phone call intervention; patient specific questions were fielded on an individual basis. This second intervention is not discussed further in this review. The letter intervention group received a standardized letter addressed to each specific patient. Both the phone call script and letter referenced the 2014 CDC immunization schedule and guidelines<br>Category and basis: PL = Personalized Letter (addressed to each patient) |
| McCaul et al, 2002<br>USA<br>n = 23,733 | Control: No reminder<br>Intervention: (1) Reminder letter from state peer review organisation (PRO), or (2) reminder letter with loss or gain frame from PRO, or (3) action letter from county public health office with date, time and place of vaccination clinics.<br>The reminder letter highlighted four main points: (a) "You should have a flu shot every year," (b) "Medicare will pay for your flu shot this fall," (c) "The flu shot is safe," and (d) "You should have your shot soon." In addition, the framing letter stated, "As a person 65 or older, you are at risk for getting a serious case of flu." The framing letter was accompanied by one of two inserts. The gain insert featured the picture and testimonial of a North Dakota woman who had received a flu shot the previous year and had not gotten the flu; the loss insert featured the picture and testimonial of another North Dakota woman who had not received a flu shot last year and had spent several days in bed, sick with the flu. More detail on the arms is provided in Chapter 4.<br>Category and basis: PL = Personalised letter (reference to age; addressed to individual; signature of doctor) |
| McDowell et al, 1986<br>Canada<br>n = 939 | Control: No reminder<br>Intervention: (1) A personal reminder by the physician, or (2) a telephone reminder by the nurse, or (3) a letter reminder. Only the latter intervention is discussed here. The letter was signed by the patient's physician and the practice nurse. The letter read: "As you know, each fall we recommend immunization against influenza for our patients who are 65 years of age or older. The vaccine is now available and if you would like to be immunized, please call to schedule an appointment."<br>Category and basis: PL = Personalised Letter (addressee selected by age, signed by physician and practice nurse) |
| Moran et al, 1992<br>USA<br>n = 409 | Control: Usual Care<br>Intervention: (1) Reminder letter offering free vaccination with an appointment, or (2) two sequential reminder letters, offering the same. The sequential reminder intervention is not discussed further in this review. The reminder letters were written at fifth-grade reading level and emphasized that: 1) immunization was medically indicated, 2) immunization did not cause influenza, 3) immunization could result in minor side effects, and 4) immunization was free and available without an appointment.<br>Category and basis: PL = Personalised Letter (advising high risk patient that immunisation is medically indicated) |
| Mullooly et al, 1987<br>USA<br>n = 2217 | Control: Did not receive the mailed cue.<br>Intervention: Personalized letter stressing the importance of influenza vaccination for high-risk elderly individuals who had been hospitalized during the past year. It was explained that immunization could help to avoid serious complications from the bout of flu and that the CDC and their personal Kaiser Permanente doctors recommend that they get a flu shot each year. Information about how and where to obtain a vaccination was also provided.<br>Category and basis: PL = Personalised Letter (described as personalised by author, letter also makes reference to people discharged from hospital in last year) |
| Nexøe et al, 1997<br>Denmark<br>n = 585 | Control: No letter<br>Intervention: (1) Patients were invited for vaccination and had to pay the GP's usual fee, or (2) patients were invited for free vaccination. The second intervention is not discussed further in this review.<br>Category and basis: PL = Personalised Letter. Letter included patient's name and GP's signature in print. |
| Roca et al, 2012<br>Spain<br>n = 2402 | Control: No intervention<br>Intervention: A personalized letter including basic information about the clinical manifestations and possible complications of influenza, and about the efficacy of the vaccine to prevent the disease, according to recommendations of the Centers for Disease Control and Prevention and the local authorities of the Comunidad Valenciana. The letter addressed common concerns about the flu shot and was written in easy-to-understand language.<br>Category and basis: PL = Personalized Letter (described as personalized by author, paper also makes reference to where patients' postal addresses were obtained from) |
| Satterthwaite et al, 1997<br>New Zealand<br>n = 2791 | Control: No reminder<br>Intervention: (1) Personalised letter recommending vaccination, or (2) personalised letter recommending visit to receive vaccine at no charge. Both letters were signed by principal. The second intervention is not discussed further in this review.<br>Category and basis: PL = Personalised Letter. |
| Terrell-Perica et al, 2001<br>USA<br>n = 6528 | Control: No letter. During the study period, the State of Hawaii Department of Health conducted routine promotional activities for influenza immunization, including press releases, immunization clinics held at pharmacies and retail stores, and health education at a large annual senior fair. In addition, pneumococcal education kits produced by the National Institute on Aging were mailed to physicians.<br>Intervention: (1) A letter encouraging recipients to take advantage of their new Medicare benefits to receive influenza immunization, or (2) a letter encouraging them to take advantage of their new Medicare benefits to receive influenza *and* pneumococcal immunizations – this intervention is not discussed further in this review. The one-page influenza immunization reminder letter was formatted in an easy-to-read, 14-point font with two prominent bullets: “Have you had your FLU shot this year?” and “Medicare covers FLU shots!”<br>Category and basis: PL = Personalized Letter (did not apply to all households, new Medicare members) |
| Yokum et al, 2018<br>USA<br>n = 228,000 | Control: No letter<br>Intervention: (1) A letter with vaccination information + picture of National Vaccine Program Officer, or (2) a letter with vaccination information + picture of Acting US Surgeon General, or (3) a letter with implementation intention prompt + picture of Acting US Surgeon General, or (4) a letter with enhanced active choice implementation prompt + picture of Acting US Surgeon General (more details in Chapter 4).<br>Category and basis: PL = Personalised Letter (addressed to recipient’s first name) |
| CDC 1995<br>USA<br>n = 190,000 | Control: No letter. Measures to increase influenza vaccination coverage including public service announcements and notices to health-care providers<br>Intervention: (1) A personalized letter and informational brochure from the Montana-Wyoming Foundation for Medical Care (MWFMC) medical director encouraging vaccination, or (2) a form letter and informational brochure from the MWFMC encouraging vaccination. |
|  | Category and basis: (1) PL + B, (2) GL + B = Personalised Letter and Brochure, and Generic Letter and Brochure (described as such by authors, also from named medical director) |
| Minor et al, 2010<br>USA<br>n = 1371 | Control: Standard clinical practice<br>Intervention: (1) A letter addressed from the clinic and signed by the clinic pharmacist and physician medical director and a copy of the Centers for Disease Control and Prevention (CDC) Influenza Vaccine Information Statement, or (2) a telephone reminder. The latter is not discussed further in this review.<br>Category and basis: PL + B = Personalised Letter and Brochure (letter signed by the clinic pharmacist and physician medical director) |
| Brimberry et al, 1988<br>USA<br>n = 787 | Control: No reminder<br>Intervention: (1) Patients received a reminder letter or, (2) a personal telephone reminder. The second intervention is not discussed further in this review. The letter emphasized that, because of “certain medical problems (for example, diabetes or heart disease),” influenza can be a serious threat to health, and that the patient’s physician had recommended that the patient be vaccinated. As a form letter was used, each patient’s personal diagnosis could not be mentioned, and the signature of a designated “influenza vaccination director” was used because of the difficulty of obtaining the signature of each patient’s personal physician. To make the vaccination convenient for the patient, no appointment was necessary, and the patient was informed of the cost.<br>Category and basis: GL = Generic Letter. |
| Buchner et al, 1987<br>USA<br>n = 655 | Control: No reminder<br>Intervention: Postcard reminder; short message on 3-inch by 5-inch card, mailed in business envelope with physician’s return address; message indicated flu season was coming, some people are at greater risk for influenza and complications, flu shots can decrease risks with minimal side effects, and it is needed each year; also provided instructions for where to obtain flu shots. By having the physician sign the cue and by using the physician’s business envelope, the cue emphasised that the physician recommended the flu shot.<br>Category and basis: PP = Personalised Postcard (signed by physician) |
| Puech et al, 1998<br>Australia<br>n = 325 | Control: Usual care, considered to be an ad hoc approach, influenced by news coverage of potential epidemics, media campaigns by vaccine manufacturers, opportunistic reminders and other secular events.<br>Intervention: A postcard encouraging patients to attend the practice for an influenza vaccination before the end of the month. The postcard stressed the seriousness of influenza as opposed to the effectiveness and safety of influenza vaccine; it also gave |
|  | availability and cost information. For ease of reading, the postcard was large (A5 format) and had clear, black-on-white large print. The postcard had a Flesch readability score of 68,14 requiring a minimum IQ of 90 to understand it (75% of the general population would understand it). Postcards had the practice logo and were mailed in a handwritten, personally addressed envelope also printed with the practice logo.<br>Category and basis: PP = Personalised Postcard (personally addressed envelope) |
| Spaulding et al, 1991<br>USA<br>n = 1068 | Control: No postcard and received routine care.<br>Intervention: A reminder postcard advising patients that their physician had determined that they were at high risk of complications should they catch the “flu,” and strongly urging them to come to the Family Practice Clinic for immunization.<br>Category and basis: PP = Personalised Postcard (the letter stated that their physician had determined that they were at high risk of complications should they catch the flu) |
| Clayton et al, 1999<br>USA<br>n = 5278 | Control: Standard member educational materials sent by mail.<br>Intervention: Postcard reminder mailed in addition to standard materials.<br>Category and basis: P = Postcard (unclear if generic or personalised as no information given in paper) |
| Larson et al, 1982<br>USA<br>n = 395 | Control: No reminder<br>Intervention: (1) Patients sent a neutral postcard mentioned influenza vaccine now available; listed telephone number for nurse appointments; addressed to “Dear Patient”; or (2) health belief model postcard, emphasizing severity of influenza, susceptibility of at-risk persons to influenza, and benefits of vaccination; addressed to “Dear Patient”, or (3) personal postcard; addressed to patient’s name and signed by clinician; postcard mentioned that influenza season is approaching and recommended the patient come in for flu shot; it listed telephone number to call and make appointment with nurse (more details in Chapter 4 and Appendix).<br>Category and basis: (1) GP = Generic Postcard (2) HBP = Health Belief Model Postcard (3) PP = Personalised Postcard |
| Moran et al, 1996<br>USA | Control: No intervention<br>Intervention: (1) A large print, illustrated educational brochure emphasizing factors important to patients in making a decision about influenza immunization, or (2) a lottery-type incentive announcing that all patients receiving influenza immunization |
| n = 797 | would be eligible for grocery gift certificates, or (3) both educational brochure and incentive.<br>Category and basis: (1) GEB; (2) Lottery; (3) GEB + Lottery. Generic Educational Brochure and financial incentive (gift certificate lottery) |

## Notes

### Competing Interest Statement

The authors have declared no competing interest.

### Funding Statement

This research was not funded.

### Author Declarations

This is a meta-analysis of previously published primary studies. Therefore, RB/oversight body approval was not required.

