## Supplemental Figures for "A Meta-Analysis of Influenza Vaccination Following Correspondence: Considerations for COVID-19"

Online supplementary file

**Section 1:** *Fig. S.1. Risk of Bias Assessment*

**Section 2:** *Fig. S.2. Funnel Plot of Log Risk Ratio for Vaccination in Participants Who Received a Messaging Intervention Relative to Control versus Standard Error*

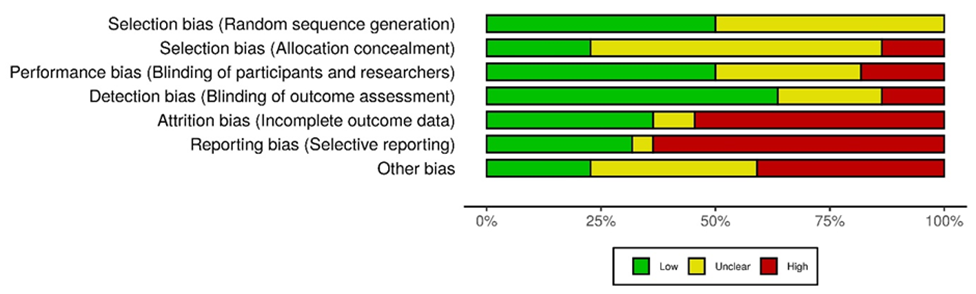

*Fig. S.1.* Risk of Bias Assessment

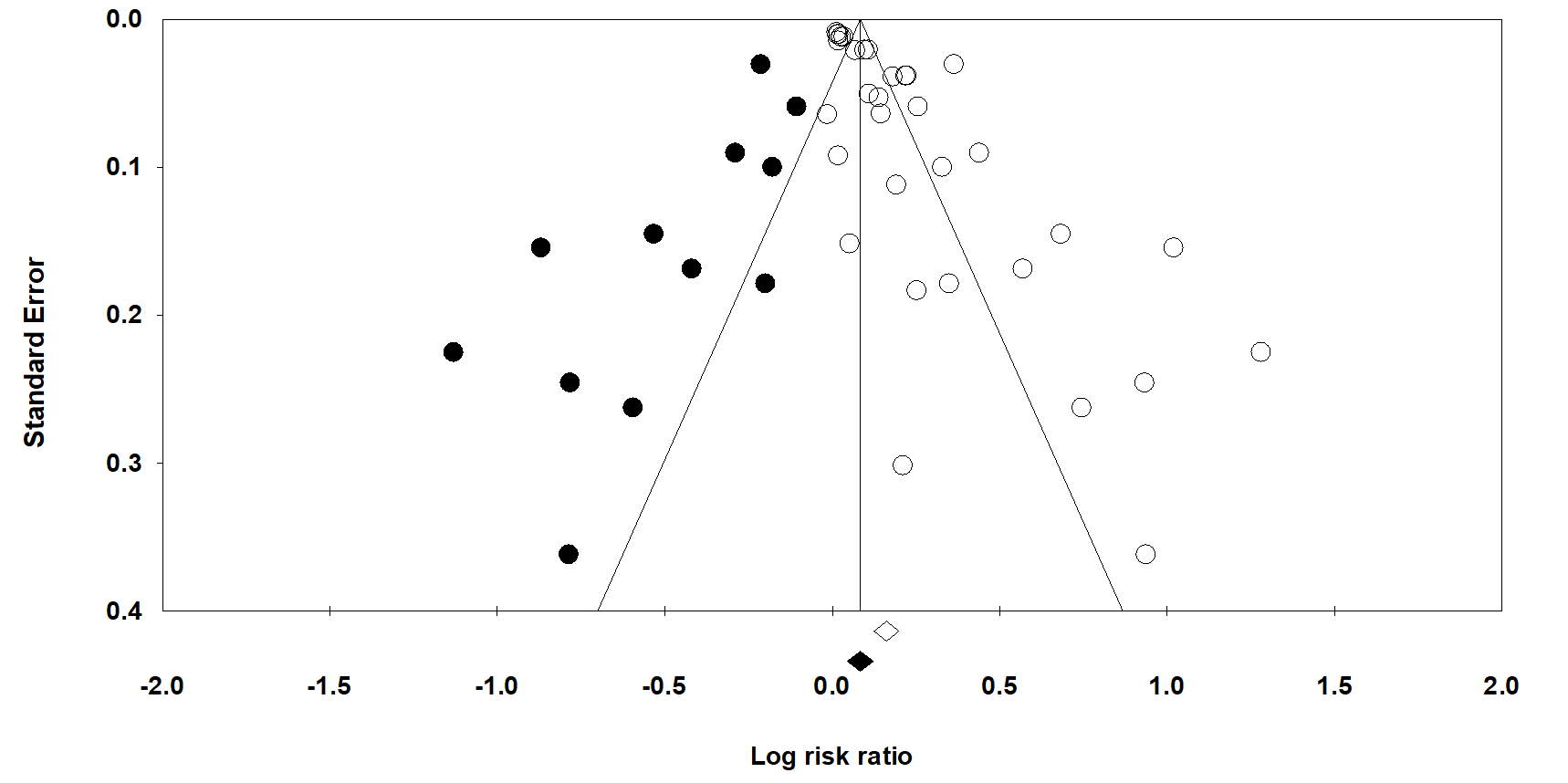

*Fig. S.2. Funnel Plot of Log Risk Ratio for Vaccination in Participants Who Received a Messaging Intervention Relative to Control versus Standard Error*

*Note.* The vertical line through the funnel plot indicates the weighted, mean effect size estimate. Clear circles indicate each of the meta-analysed studies. Black circles indicate the imputed 12 hypothetical, missing studies following trim and fill analysis. The clear diamond corresponds to the log of the observed, weighted, mean effect size estimate (RR = 1.18). The black diamond corresponds to the log of the adjusted, weighted, mean effect size estimate following imputation (RR = 1.09).
